# Revolutionizing Postoperative Ileus Monitoring: Exploring GRU-D’s Real-Time Capabilities and Cross-Hospital Transferability

**DOI:** 10.1101/2024.04.24.24306295

**Authors:** Xiaoyang Ruan, Sunyang Fu, Heling Jia, Kellie L. Mathis, Cornelius A. Thiels, Patrick M. Wilson, Curtis B. Storlie, Hongfang Liu

## Abstract

**Background:** Postoperative ileus (POI) after colorectal surgery leads to increased morbidity, costs, and hospital stays. Identifying POI risk for early intervention is important for improving surgical outcomes especially given the increasing trend towards early discharge after surgery. While existing studies have assessed POI risk with regression models, the role of deep learning’s remains unexplored.

**Methods:** We assessed the performance and transferability (brutal force/instance/parameter transfer) of Gated Recurrent Unit with Decay (GRU-D), a longitudinal deep learning architecture, for real-time risk assessment of POI among 7,349 colorectal surgeries performed across three hospital sites operated by Mayo Clinic with two electronic health records (EHR) systems. The results were compared with atemporal models on a panel of benchmark metrics.

**Results:** GRU-D exhibits robust transferability across different EHR systems and hospital sites, showing enhanced performance by integrating new measurements, even amid the extreme sparsity of real-world longitudinal data. On average, for labs, vitals, and assisted living status, 72.2%, 26.9%, and 49.3% respectively lack measurements within 24 hours after surgery. Over the follow-up period with 4-hour intervals, 98.7%, 84%, and 95.8% of data points are missing, respectively. A maximum of 5% decrease in AUROC was observed in brutal-force transfer between different EHR systems with non-overlapping surgery date frames. Multi-source instance transfer witnessed the best performance, with a maximum of 2.6% improvement in AUROC over local learning. The significant benefit, however, lies in the reduction of variance (a maximum of 86% decrease). The GRU-D model’s performance mainly depends on the prediction task’s difficulty, especially the case prevalence rate. Whereas the impact of training data and transfer strategy is less crucial, underscoring the challenge of effectively leveraging transfer learning for rare outcomes. While atemporal Logit models show notably superior performance at certain pre-surgical points, their performance fluctuate significantly and generally underperform GRU-D in post-surgical hours.

**Conclusion:** GRU-D demonstrated robust transferability across EHR systems and hospital sites with highly sparse real-world EHR data. Further research on built-in explainability for meaningful intervention would be highly valuable for its integration into clinical practice.

## Background

Postoperative ileus (POI) poses a common challenge following colorectal surgery, contributing to heightened morbidity, increased costs, and prolonged hospital stays ^1^. With an occurrence rate of 10%-30% and typically manifesting within 6-8 days post-surgery ^2^, addressing POI has gained significance for hospitals due to various reasons. These include a payment reform that emphasizes extended care episodes and Medicare penalties associated with readmissions within 30 days, an increasing preference for outpatient surgery to conserve limited hospital resources amid the COVID-19 pandemic ^3^, along with a contentious surge in interest for same-day or next-day discharge ^4^. Efficiently identifying the risk of POI to enable early intervention stands as a crucial factor in enhancing surgical outcomes. While various studies have discussed the prediction of POI ^5^ ^6^ ^7^ ^8^ ^9^ ^10^, predominantly utilizing multivariate logistic regression techniques, there exists a notable gap in research concerning deep learning-based approaches in POI prediction. This is particularly intriguing given the widespread application of deep learning in tackling other postoperative complications ^11 12 13^.

We previously benchmarked the Gated Recurrent Unit with Decay (GRU-D), a RNN based architecture proposed by Che et al ^14^, in making real-time risk assessment of post-surgical complications including superficial infection, wound infection, organ space infection, and bleeding ^12^. GRU-D exhibited advantageous attributes including automated missing imputation, the flexibility to tailor sampling intervals, and the capacity to update and enhance risk assessment by incorporating newly received measurements. These features position GRU-D as an ideal candidate for risk modeling associated with longitudinal clinical data, a domain often characterized by substantial missings in data, asynchronous updates, and the need for prompt risk assessment.

The considerable practical values that GRU-D holds underscores the importance of examining its applicability across a variety of real-world contexts. Here we aim to assess the feasibility, performance, and transferability of GRU-D in risk modeling of POI across two electronic health record (EHR) systems in three separate hospital sites affiliated with Mayo Clinic. The results are compared with atemporal logistic regression and random forest models on both pre-surgical and post-surgical periods to evaluate the strengths and limitations of GRU-D based strategy in POI risk assessment. This evaluation takes place against the backdrop of the increasing significance of time series data, a prevalent format in clinical settings used to chronicle patient longitudinal information, within the realm of transferability research. Notably, prior research on transferability involving time series data has primarily centered around neurology and cardiology ^15^ ^16^. There exists a substantial dearth of research concerning the transfer learning of time series data related to surgical contexts, with only a limited number of studies being documented ^17^ ^18^.

## Materials and Methods

### CRC surgery samples

The study consists of 7349 colorectal surgery records from 7103 patients with colorectal surgeries performed at Mayo Clinic Rochester (MR), Arizona (MA), Florida (MF) hospital sites between 2006 and 2022 as part of the National Surgical Quality Improvement Program (NSQIP) cohort ^19^. Specifically, due to a transition from Centricity to EPIC system in 2018, data from MR is split into two parts as MR centricity (*MR_cc_* ) and MR EPIC (*MR_ep_*). Whereas data from both MA and MF are after 2018 with the EPIC system. The distribution of surgery time by sites and EHR systems are illustrated in Fig 1. The baseline characteristics of the patients are shown in Table 1. The study was approved by the Mayo Clinic institutional review board (IRB number: 15-000105). All patients included in this study had consent for their data to be used for research.

**Fig 1.**
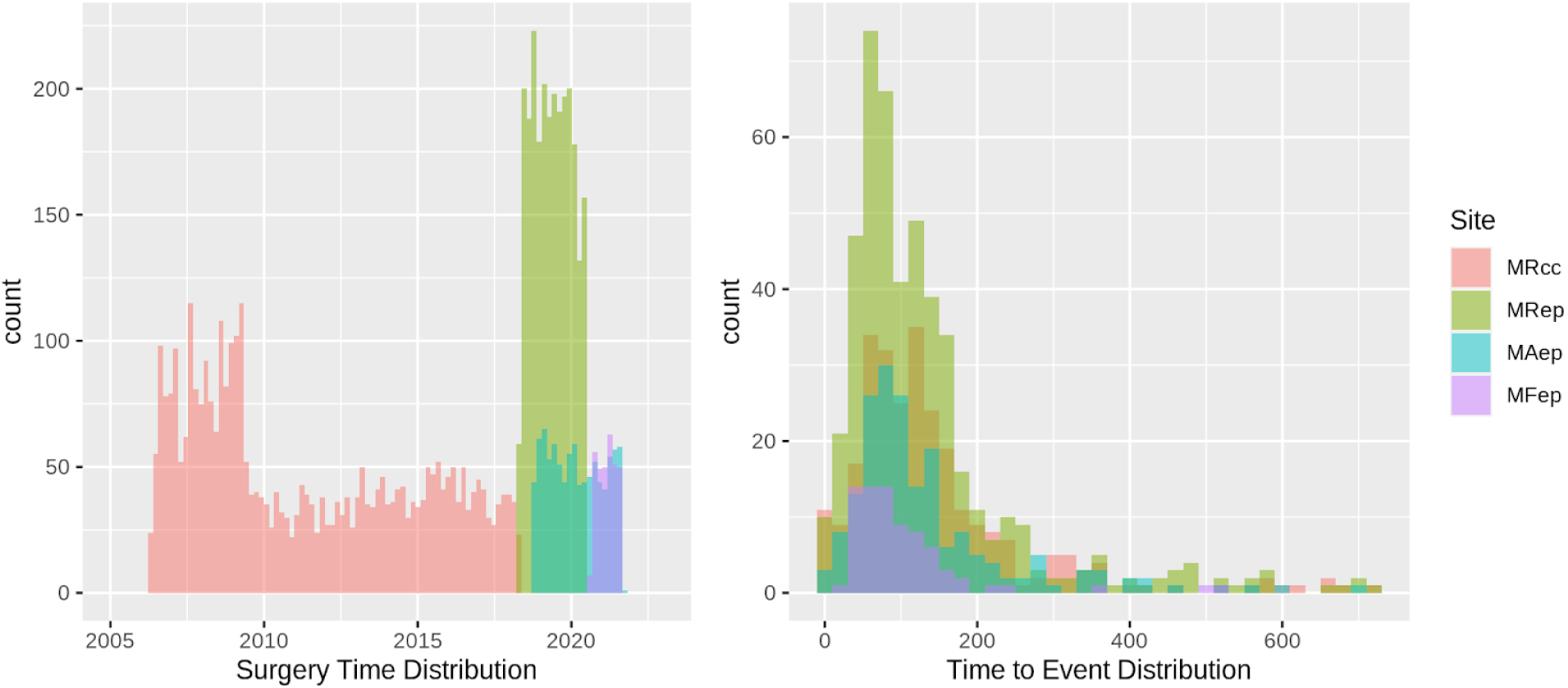
Distribution of colorectal surgery time (left) and time to event (right) across site-systems.

**Table 1.** Baseline characteristic of study population.

| Hospital site full name | Mayo Clinic Rochester | Mayo Clinic Rochester | Mayo Clinic Arizona | Mayo Clinic Florida |  |
| --- | --- | --- | --- | --- | --- |
| Hospital site abbreviation | MR | MR | MA | MF | ALL |
| EHR system | Centricity | EPIC | EPIC | EPIC | - |
| Time period | 2006-2018 | 2018-2021 | 2018-2022 | 2020-2022 |  |
| Site-System Abbreviation | *MR<sub>cc</sub>* | *MR<sub>ep</sub>* | *MA<sub>ep</sub>* | *MF<sub>ep</sub>* | - |
| Number of patients | 3535 | 2352 | 898 | 318 | 7103 |
| Number of records | 3598 | 2493 | 932 | 326 | 7349 |
| Age at surgery | 59(45,71)<sup>a</sup> | 58(45,69) | 61(47,71) | 62(48,71) | 59(46,70) |
| Gender |  |  |  |  |  |
| Male | 1767(50%) | 1098(47%) | 439(49%) | 147(46%) | 3451(49%) |
| Female | 1768(50%) | 1252(53%) | 459(51%) | 171(54%) | 3650(51%) |
| Ileus cases | 274(7.6%) | 475(19.1%) | 187(20.1%) | 76(23.3%) | 1012(13.8%) |
| Race |  |  |  |  |  |
| Caucasian | 3315(94%) | 2161(92%) | 818(91%) | 289(91%) | 6583(93%) |
| Non-caucasian | 220(6%) | 191(8%) | 80(9%) | 29(9%) | 520(7%) |
| Surgery subtype |  |  |  |  |  |
| Laparoscopy, colectomy | 978(27%) | 530(21%) | 268(29%) | 68(21%) | 1844(25%) |
| Laparoscopy, colectomy, partial | 634(18%) | 578(23%) | 265(28%) | 87(27%) | 1564(21%) |
| Removal of colon | 701(19%) | 409(16%) | 120(13%) | 55(17%) | 1285(17%) |
| Removal of colon, partial | 505(14%) | 460(18%) | 147(16%) | 69(21%) | 1181(16%) |
| Removal of rectum | 255(7%) | 111(4%) | 18(2%) | 9(3%) | 393(5%) |
| Others<sup>b</sup> | 525(15%) | 405(16%) | 124(13%) | 38(12%) | 1092(15%) |
| Surgery time duration, minutes | 188(136,256) | 238(158,353) | 198(138,286) | 236(185,319) | 204(143,290) |

<sup>a</sup> Data shown as n(%) or median(IQR)
<sup>b</sup> See Table S1 for a comprehensive list of subtypes

### Ileus status ascertainment

The identification of ileus was followed by the application of a standardized NLP pipeline through the OHNLP infrastructure. Additional details about the NLP methodology can be found in our previous studies ^20^ ^21^. Briefly, the development of the ileus algorithm followed an integrative process, which included corpus annotation, symbolic ruleset prototyping, ruleset refinement, and final evaluation. The validated ileus rulesets were then integrated into the OHNLP Backbone and MedTaggerIE NLP pipeline, which includes a built-in context classifier (e.g., status, subject, and certainty).

### Training data and held-out data selection

For surgery records of each site, 30% were randomly selected as held-out, and the remaining 70% were randomly splitted into six equal-sized training chunks (Table 2). The randomization was performed 100 times to select the one that minimizes the difference between chunk-level and site-level characteristics on ileus complication rate, surgery duration, gender, and race.

**Table 2.** Training and held-out split.

| Table 2 Training and held-out split |  |  |  |  |  |
| --- | --- | --- | --- | --- | --- |
| Site-System | $MR_{cc}$ | $MR_{ep}$ | $MF_{ep}$ | $MA_{ep}$ | $ALL$ |
| Total | 3598 | 2493 | 326 | 932 | 7349 |
| Ileus cases | 274 | 475 | 76 | 187 | 1012 |
| case (%) | 7.60% | 19.10% | 23.30% | 20.10% | 13.80% |
| Held-out | 1079 | 747 | 97 | 278 | 2201 |
| cases | 75 | 144 | 25 | 57 | 301 |
| case (%) | 7.00% | 19.30% | 25.80% | 20.50% | 13.70% |
| Training | 2519 | 1746 | 229 | 654 | 5148 |
| cases | 199 | 331 | 51 | 130 | 711 |
| case (%) | 7.90% | 19.00% | 22.30% | 19.90% | 13.80% |

### Transfer learning naming rules and transfer schemes

Throughout the paper we adhere to the following naming rules for clarity. For each dataset, subscripted *MR_cc_, MR_ep_, MF_ep_, MA_ep_* represent the dataset originated from Mayo Clinic Rochester Centricity, Rochester EPIC, Florida EPIC, and Arizona EPIC system, respectively. Source training (*S^Tr^*) and source held-out (*S^To^*) represent data from the training and held-out chunks of source site-system(s), respectively. Target training (*T^Tr^*) and target held-out (*T^Ho^*) represent data from the training and held-out chunks of the target site-system(s), respectively. E.g., 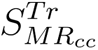 represents source training dataset from Mayo Clinic Rochester Centricity system. We use function *M*() to represent deep learning model to make a distinction from the corresponding dataset it was trained upon. E.g. 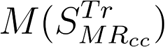 represent models trained from 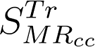 dataset.

For each site-system, the six chunks of training data were used to train six independent models by iteratively leaving one chunk out. The models were named, take *MR_cc_*for example, as 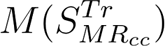 (an ensemble of six models). We consider five model training scenarios (Take *MR_ep_* for example) including local learning (i.e. training on 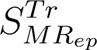 and predict 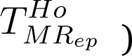 and four scenarios of transfer learning (transfer from 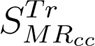 for example) detailed below, according to solution based categorization ^22^.

***Brutal-force transfer:*** Directly apply 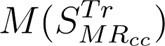 to 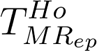.

***Single-source instance transfer:*** Combine training data records from *MR_cc_* and *MR_ep_* and apply 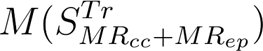 to 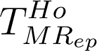 .

***Parameter transfer:*** Continuous training of 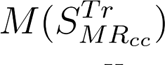 on *MR_ep_*. The models were named 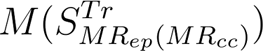 , which were then applied to 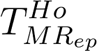 .

***Multi-source instance transfer:*** Train brand new models 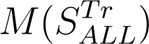 by combining training data from all sites and then applied to 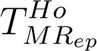 .

### GRU-D model architectures

The basic architecture of the GRU-D model was described by ^23^, here we recapitulate the equations for handling missing values.

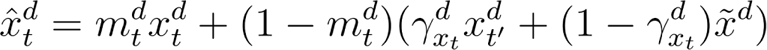

where 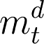 is the missing value indicator for feature *d* at timestep *t*. 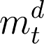 takes value 1 when 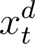 is observed, or 0 otherwise, in which case the function resorts to weighted sum of the last observed value 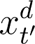 and empirical mean 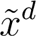 calculated from the training data for the *d* th feature. Furthermore, the weighting factor 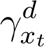 is determined by

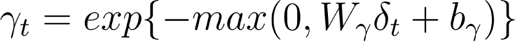

where *W_γ_* is a trainable weights matrix and *δ_t_* is the time interval from the last observation to the current timestep. When *δ_t_* is large (i.e. the last observation is far away from current timestep), *γ_t_* is small, results in smaller weights on the last observed value 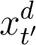 , and higher weights on the empirical mean 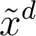 (i.e. decay to mean).

For each timestep of each patient, sigmoid activation function is applied to the hidden output to generate a predicted probability *y_it_* (0∼1) of developing ileus within 30 days after surgery. *y_it_* is evaluated against the true class label (*◯*), i.e., whether or not the patient actually developed ileus within 30 days of surgery. Specifically, the loss function is as follow

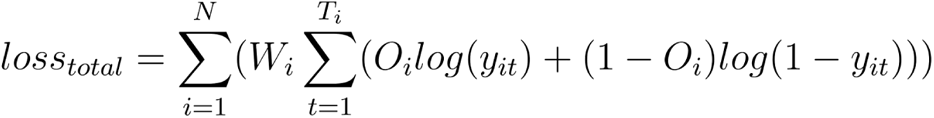

where *N* is the number of training samples in a batch. *W_i_* is the weights for sample *i* (minor group inverse weighted). *T_i_* is the maximum allowed timesteps (i.e. the timesteps before ileus onset, if any) for sample *i*. *◯_i_* is the observed outcome. *y_it_* is the predicted probability for sample *i* at timestep *t*.

### Competing models

The performance of GRU-D model was compared with atemporal logistic regression with lasso regularization (Logit) (R package *glmnet*) and atemporal random forest (RF) (R package *randomForestSRC*). For both models, we implemented unlimited time last value carry forward to fill in the missing values. The remaining missing values were imputed with RF adaptive tree imputation ^24^ with three iterations. Specifically, the imputation for training and held-out data from different sites were implemented separately. Separate models were trained at integer days (i.e., -4, -3 … 13, 14) and used to make predictions at corresponding time points. Features with missing proportions greater than 0.2 were removed from analysis. For the Logit model, the default configurations (alpha=1, nlambda=100) were adopted. For the RF model, the number of trees were set to 100 with a minimum of 10 terminal nodes.

### Input features and feature selection

We included features that appeared in all four site-systems (Table S1). Notably, for the GRU-D model no exclusion of features by missing proportion was performed, to respect real world scenarios where data availability may vary across sites.

We meticulously reviewed each feature to classify it as continuous or categorical. To support transfer learning, we manually established a biologically meaningful mapping from Centricity to EPIC categories for each categorical feature, resulting in a semi-homogeneous transfer learning setup. For example, we mapped the 29 fine-grained “Diet” categories in the Centricity system to the 3 corresponding categories (“Nothing by mouth,“ “Diet 50,” and “Diet 100”) in the EPIC system.

Categorical features underwent one hot encoding based on categories presented across all four site-systems, while continuous features were z-score normalized using mean and standard deviation values calculated from the source training data. During parameter transfer learning, we updated the mean and standard deviation values with corresponding values from the target training data, if available. To avoid extreme values, the z-score values were hard clamped to between -5 and 5 before being fed into the machine learning models.

### Sampling scheme of dynamic and static features

Dynamic features were sampled at a 4-hour interval from 4 days before surgery to 14 days after surgery, using the last measurement in the past 4 hours, or marked as missing if no value was spotted. This generates 109 timesteps for each feature of each patient. Static features were replicated across time. Age is converted to within range 0∼1 by dividing by 100. Surgery time duration is hourly based. Surgery subtypes are one hot encoded into 14 categories.

### Evaluation metrics

We benchmarked the models’ performance with established metrics such as area under the receiver operating characteristic (AUROC), average precision, the optimal cutoff threshold (which maximizes the geometric mean of sensitivity and specificity), and the F-score (at the optimal threshold). Furthermore, we also experimented with a panel of customized metrics focused on clinical utility. These include setting up a monitoring ward specifically for the highest risk quartile (i.e., the fourth quartile) and record the number of unique ileus cases identified in each time step (referred to as Q4-cases), the cumulative number of unique Q4-cases over the entire follow-up period (Q4-cases accum), and the percentage of accumulated cases among all monitored patients (Accum Case %, i.e. accumulated precision). We also presented for each model the flag rate (i.e. the proportion of patients that need to be monitored) and positive predictive value (PPV) in order to achieve 60% sensitivity throughout each time point of the follow-up. To ensure comparability between GRU-D and atemporal models, the clinical utility related metrics were evaluated exclusively on integer days (-96h, -72h, …, 24h, 48h…). To ensure the accuracy of our evaluation metrics, we removed patients who had already developed ileus complications before the specific timestep of interest, thus eliminating confounding effects from prior occurrences of the complication.

We examined two approaches for assessing metrics in our study. Taking AUROC as an example, the first approach, termed timestep-specific analysis (AUROC_t_), involves evaluating the predicted probability at a particular timestep *t* against ileus outcome, while excluding patients with ileus diagnosed prior to that timestep. The second approach, termed last-timestep analysis (AUROC_last_), evaluates the predicted probability at the final timestep before ileus onset (or the 109th timestep if no ileus) against ileus outcome. This second approach is specifically designed to assess the transferability of GRU-D models using all available information. In our study, we evaluated AUROC, average precision, and F-score using both strategies, whereas the remaining metrics were only evaluated using the first strategy.

### Permutation feature importance

The permutation feature importance test is performed by randomly rearranging the sample IDs of a given feature in the target dataset. For each of the six models (obtained from the six-fold cross-validation) five permutations were executed, resulting in a total of 30 permutations for each feature at every timestep. The AUROC_t_ was assessed both before and after the permutation. The mean discrepancy in AUROC_t_ across all timesteps (calculated as before permutation minus after permutation) was used to gauge the model’s reliance on the specific feature.

## Results

### Feature sparsity of real-world data

To demonstrate the level of feature sparsity derived from real-world EHR data, we visualized the missing proportion of selected features (Labs, Vitals, ADL (Assisted Living Status)) in Fig 2, and CCS codes in Fig S3. The EHR data from the studied hospital sites is predominantly sparse across multiple modalities throughout the duration of follow-up. From surgery completion (index date) to within 24 hours after surgery, there are on average 72.2% labs, 26.9% vitals, 49.3% ADL, 95.4% CCS codes lacking a measurement. With a standardized 4 hour interval based time grid, the average missing proportions throughout the entire follow-up are 98.7%, 84%, 95.8%, and 95.5% for labs, vitals, ADL, and CCS codes, respectively. Notably, in adhering to a realistic approach regarding data availability, no feature was eliminated based on its missing proportion in the assessment of GRU-D models. Consequently, the extent of data sparsity depicted in Fig 2 and S3 reflects the input consumed by the GRU-D models.

**Fig 2.**
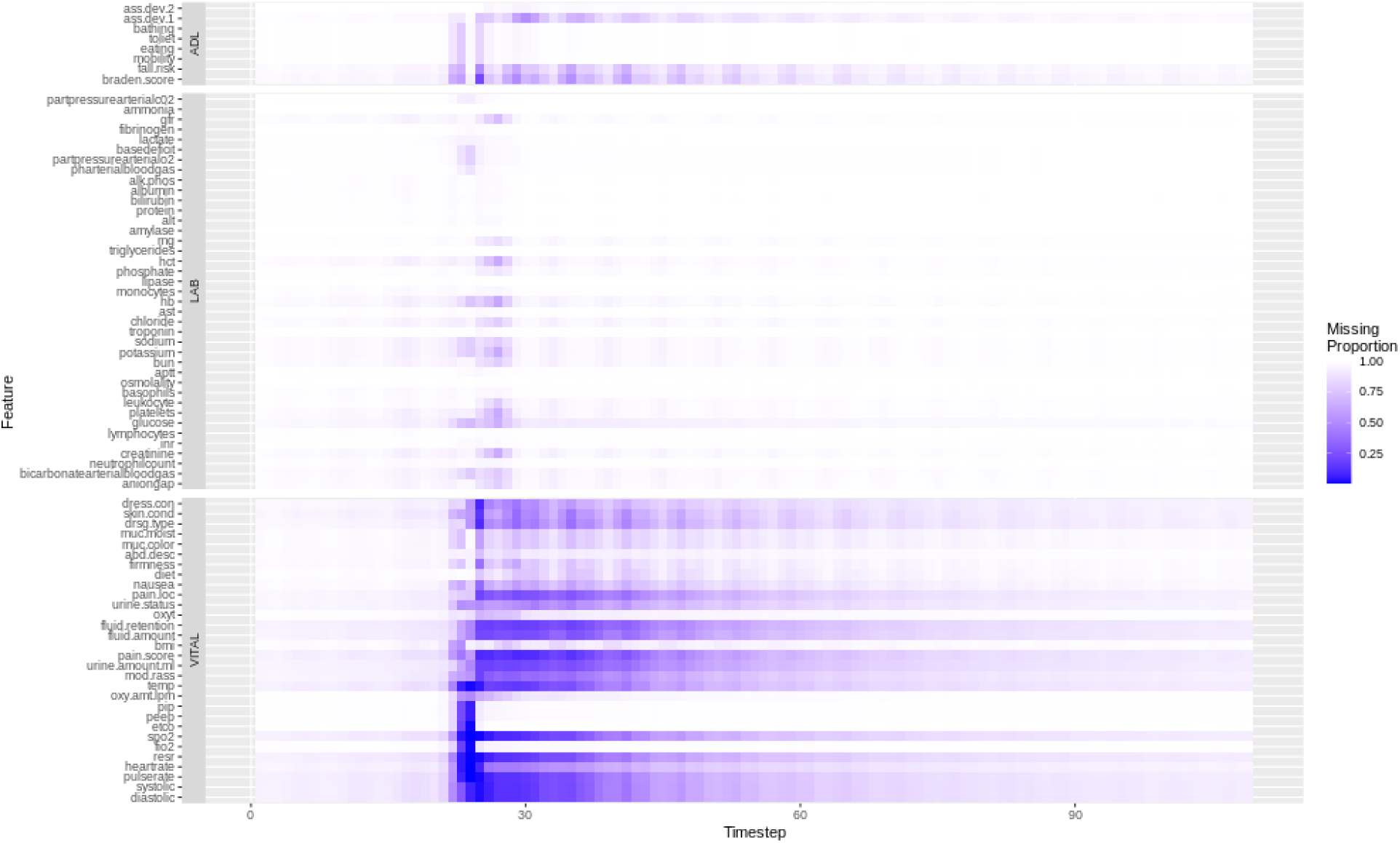
Missing proportion of selected features throughout each timesteps of follow-up, from 4 days before surgery to 14 days after surgery.

### GRU-D Transferability based on last timestep analysis

#### Multi-source instance transfer has overall the best performance

After investigating the transferability between different EHR systems within the same hospital site, and same EHR system between different hospital sites, a general observation is that multi-source instance transfer with data source indicator has overall the best performance (Table 3,4,5 *ALL (w)ds*), slightly surpassing without data source indicator (Table 3,4,5 *ALL (w/o)ds*). However, the enhancement compared to local learning is only marginal. E.g. The improvement in AUROC_last_ ranges between a minimum of 0.4% (Table 3 *MR_ep_*0.935 vs *ALL (w)ds* 0.939) and a maximum of 2.6% (Table 4 *MF_ep_*0.883 vs *ALL (w)ds* 0.906). Whereas the significant benefit lies in the reduction of variance. E.g. For the EPIC site (*MF_ep_*) with the smallest training sample size (Table 2, n=226), the CI of AUROC_last_ reduced 86%, from 0.042 on local learning to 0.006 on multi-source instance transfer (Table 4). Similarly, the CI of AvgPrec_last_ reduced 56% from 0.071 to 0.031.

**Table 3.** Centricity to EPIC transferability within MR site.

| | Training<br>Source<br>data ($S^T$) | $MR_{cc}$ | $MR_{ep}$ | $MR_{cc} +$<br>$MR_{ep}$<br>(w/o)$d$<br>$S^a$ | $MR_{cc} +$<br>$MR_{ep}$<br>(w)$ds^b$ | $MR_{ep}$<br>($MR_{cc}$) | $MR_{cc}$<br>($MR_{ep}$) | $ALL$<br>(w/o)$ds$ | $ALL$<br>(w)$ds$ |
| --- | --- | --- | --- | --- | --- | --- | --- | --- | --- |
| $T_{MR_{ep}}^{Ho}$ | AUROC<sub>last</sub> | 0.882<br>,<br>0.022 | 0.935,<br>0.006 | 0.925,<br>0.01 | 0.929,<br>0.002 | 0.929,<br>0.013 | 0.907,<br>0.014 | 0.935,<br>0.006 | **0.939,**<br>**0.003** |
|  | AvgPrec<sub>last</sub> | 0.601<br>,<br>0.056 | 0.704,<br>0.038 | 0.683,<br>0.028 | 0.707,<br>0.024 | 0.691,<br>0.040 | 0.642,<br>0.035 | 0.709,<br>0.032 | **0.72,**<br>**0.028** |
|  | F-score<sub>last</sub> | 0.643<br>,<br>0.047 | 0.73,<br>0.009 | 0.7,<br>0.023 | 0.709,<br>0.009 | 0.728,<br>0.022 | 0.679,<br>0.026 | **0.737,**<br>**0.014** | 0.736,<br>0.014 |
| $T_{MR_{cc}}^{Ho}$ | AUROC<sub>last</sub> | 0.886<br>,<br>0.009 | 0.888,<br>0.011 | 0.882,<br>0.018 | 0.883,<br>0.016 | **0.9,**<br>**0.009** | 0.89,<br>0.01 | 0.89,<br>0.012 | 0.893,<br>0.009 |
|  | AvgPrec<sub>last</sub> | 0.369<br>,<br>0.016 | **0.395,**<br>**0.018** | 0.331,<br>0.043 | 0.358,<br>0.041 | 0.393,<br>0.018 | 0.342,<br>0.038 | 0.372,<br>0.024 | 0.366,<br>0.05 |
|  | F-score<sub>last</sub> | 0.432<br>,<br>0.014 | **0.463,**<br>**0.03** | 0.411,<br>0.048 | 0.428,<br>0.034 | 0.465,<br>0.031 | 0.403<br>0.032 | 0.446,<br>0.033 | 0.438,<br>0.032 |

**Table 4.** EPIC transferability from MR to MF site.

| | Training Source data ($S^T$) | $MR_{ep}$ | $MF_{ep}$ | $MR_{ep} + MF_{ep}$ (w/o)ds<sup>a</sup> | $MR_{ep} + MF_{ep}$ (w)ds<sup>b</sup> | $MF_{ep}$ ($MR_{ep}$) | ALL (w/o)ds | ALL (w)ds |
| --- | --- | --- | --- | --- | --- | --- | --- | --- |
| $T_{MF_{ep}}^{Ho}$ | AUROC<sub>last</sub> | 0.904, 0.007 | 0.883, 0.042 | 0.903, 0.005, | 0.899, 0.01 | 0.906, 0.009 | 0.895, 0.01 | **0.906, 0.006** |
|  | AvgPrec<sub>last</sub> | 0.652, 0.026 | 0.652, 0.071 | 0.635, 0.015 | 0.668, 0.032 | 0.657, 0.024 | 0.648, 0.045 | **0.691, 0.031** |
|  | F-score<sub>last</sub> | 0.804, 0.022 | 0.745, 0.064 | 0.813, 0.014 | 0.777, 0.022 | 0.807, 0.02 | 0.792, 0.033 | **0.815, 0.015** |

**Table 5.** EPIC transferability from MR to MA site.

| | Training Source data ($S^T$) | $MR_{ep}$ | $MA_{ep}$ | $MR_{ep} + MA_{ep}$ (w/o)ds<sup>a</sup> | $MR_{ep} + MA_{ep}$ (w)ds<sup>b</sup> | $MA_{ep}$ ($MR_{ep}$) | ALL (w/o)ds | ALL (w)ds |
| --- | --- | --- | --- | --- | --- | --- | --- | --- |
| $T_{MA_{ep}}^{Ho}$ | AUROC<sub>last</sub> | 0.889, 0.013 | 0.894, 0.013 | 0.896, 0.011 | 0.894, 0.01 | 0.896, 0.005 | 0.903, 0.006 | **0.911, 0.009** |
|  | AvgPrec<sub>last</sub> | 0.624, 0.017 | 0.657, 0.022 | 0.650, 0.048 | 0.67, 0.029 | 0.646, 0.023 | **0.692, 0.02** | 0.675, 0.024 |
|  | F-score<sub>last</sub> | 0.726, 0.027 | 0.723, 0.013 | 0.708, 0.018 | 0.726, 0.019 | 0.722, 0.034 | **0.726, 0.019** | 0.716, 0.02 |

<sup>a</sup> without data source indicator<sup>b</sup> with data source indicator
Bold font indicate the best performed model data shown at metric, error margin of 95% CI

#### Parameter and instance transfer perform similarly with two data sources

For models trained on only two data sources, parameter transfers (Table 3 *MR_ep_(MR_cc_),* Table 4 *MF_ep_(MR_ep_)*, Table 5 *MA_ep_(MR_ep_)*) perform similarly as their single-source instance transfer counterparts, either with or without data source indicator (e.g. Table 3 *MR_cc_ + MR_ep_ (w)(w/o)ds*). Data source indicator shows mixed effect on performance when there are only two data sources. For both parameter and instance transfers, there is generally no significant enhancement over local learning either in the evaluation metric or CI, with the exception of the EPIC site (*MF_ep_*) that has the smallest training sample size (Table 4).

#### Brutal-force transfer outperform local learning in specific scenarios

For AUROC_last_, we see up to 5% decrease among all brutal-force transfers explored in this study (Table 2 *MR_ep_* source to *MR_ep_* target (0.935) vs *MR_cc_* target (0.888)). Whereas the performance of AvgPrec_last_ and F-score_last_ depends predominantly on the target dataset (Table 2 *MR_ep_*source to *MR_ep_* target (AvgPrec_last_ 0.704) vs *MR_cc_*target (AvgPrec_last_ 0.395)). While brutal-force transfer is poorer than instance or parameter transfer under most circumstances (Table 3,4,5), it outperforms local learning in scenarios including 1) the outcome prevalence in the local dataset is low. E.g. Models trained with *MR_ep_* training data have significantly better AvgPrec_last_ (0.395) and F-score_last_ (0.463) in predicting *MR_cc_* target than *MR_cc_* local learning (Table 4 AvgPrec_last_ 0.369, CI(0.353,0.385); F-score_last_ 0.432, CI(0.418,0.446)). 2) The local dataset has a small sample size. E.g, Models trained with *MR_ep_* training data have higher AUROC_last_ and F-score_last_ and remarkably lower variance than *MF_ep_* local learning (Table 4 *MR_ep_* vs *MF_ep_* on *MF_ep_* target).

#### Negative transfer

Negative transfer was only observed when transfer from Centricity to EPIC system (Table 3 *MR_cc_* + *MR_ep_* vs *MR_ep_* on *MR_ep_*target). Compared to local learning with EPIC data only, the performance of AUROC_last_, AvgPrec_last_, and F-score_last_ decreased remarkably after incorporating training data from the Centricity system, regardless of transferring strategy. However, no notable negative transfer was observed when transfer from EPIC to Centricity (Table 3 *MR_cc_* + *MR_ep_* vs *MR_cc_* on *MR_cc_* target).

### GRU-D Transferability based on timestep specific analysis

Being a dynamic time series model, GRU-D inherently provides risk predictions at each timestep during the follow-up period. Hence, evaluating the model’s transferability at the timestep level is essential. Here we made the following major observations

1. Multi-source instance transfer shows overall optimal performance on the majority of evaluation metrics both before and after index date. E.g. It significantly excels in predicting one of EPIC targets (*MA_ep_*) before index date (Fig 5). It also yields a significantly improved Flag rate/Flag PPV when forecasting the another EPIC target (*MF_ep_*) which has the smallest sample size (Fig 4).
2. Brutal-force transfer is notably suboptimal in Centricity to EPIC transfer after index date (Fig 3), but not cross site transfer within EPIC system (Fig 4,5). Notably, it exhibits a generally higher variance a few days post-index date as the count of remaining cases decreases (Fig 3,5).
3. Local learning is only marginally poorer than other transfer strategies on certain metrics (e.g. Fig 3 Q4-cases, AvgPrec). The only scenario where local learning significantly underperforms is in predicting one EPIC target (*MA_ep_*) on AUROC and Flag rate a few days post-index date (Fig 5).
4. The model performance is predominantly determined by the prediction task, whereas the training data plays a very limited role. This is evident from the observation that when the models trained with Centricity training data (*MR_cc_*) are brutal-force transferred to EPIC target (*MR_ep_*) , they exhibit significantly better performance than being applied locally (i.e. to MRcc target)(Supp Fig 1).
5. The marginal improvement in traditional metrics like AUROC, precision, and F1-score does not necessarily translate to improvement in more clinical utility oriented metrics like accumulated Q4 cases and accumulated case percentage (Fig 3,4)

**Fig 3.**
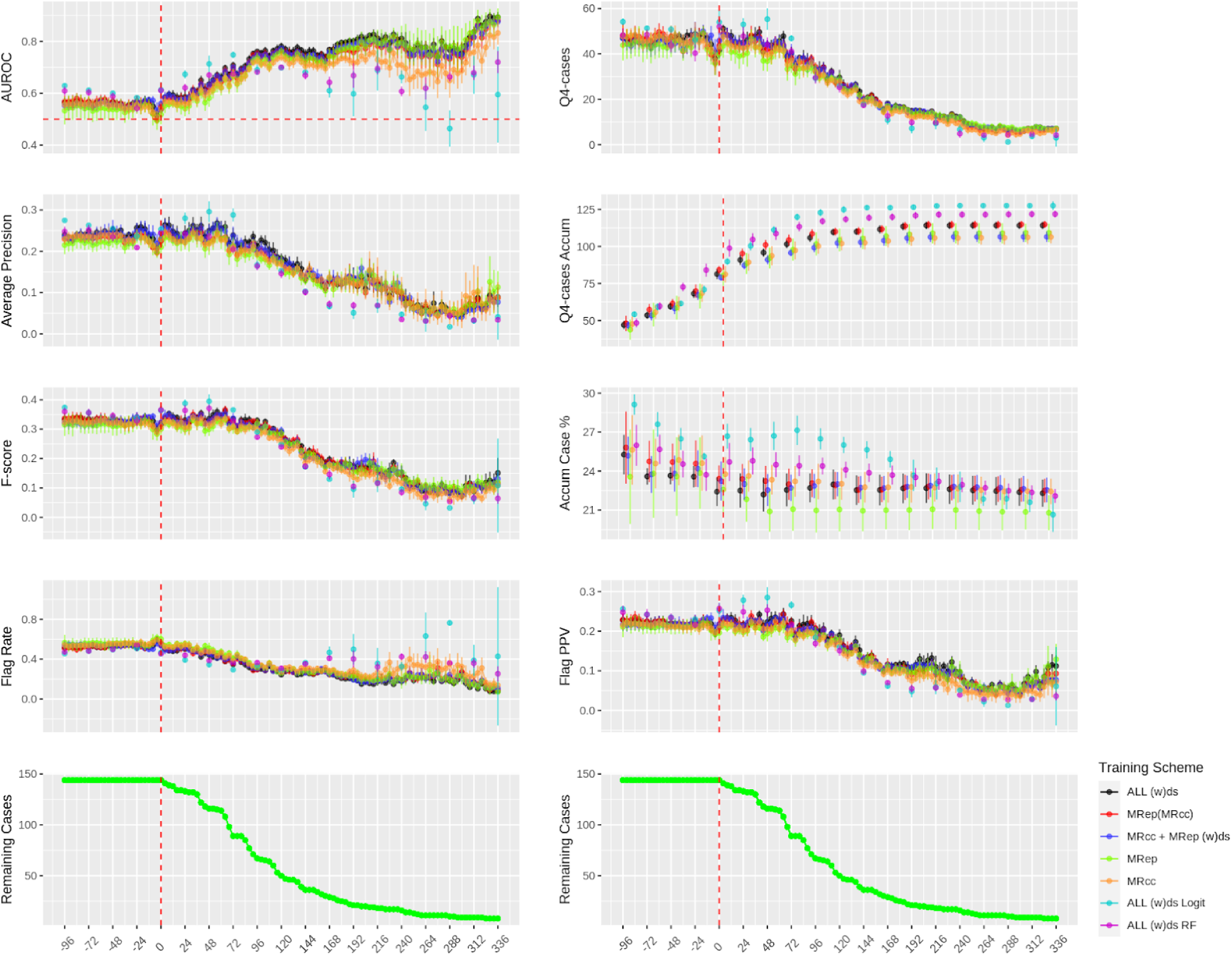
Centricity to EPIC transferability evaluated by timestep-specific analysis. All predictions made on 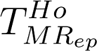 . **ALL**: instance transfer from 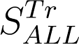 ; **MR_ep_(MR_cc_)**: parameter transfer from 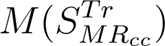 ; **MR_cc_ + MR_ep_**: instance transfer from *MR_cc_*; **MR_ep_**: local learning; **MR_cc_**: brutal-force transfer from 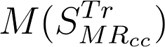 ; **ALL w(ds) Logit**: Logit based on 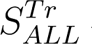 with data source indicator; **ALL w(ds) RF:** Random forest based on 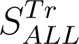 with data source indicator

**Fig 4.**
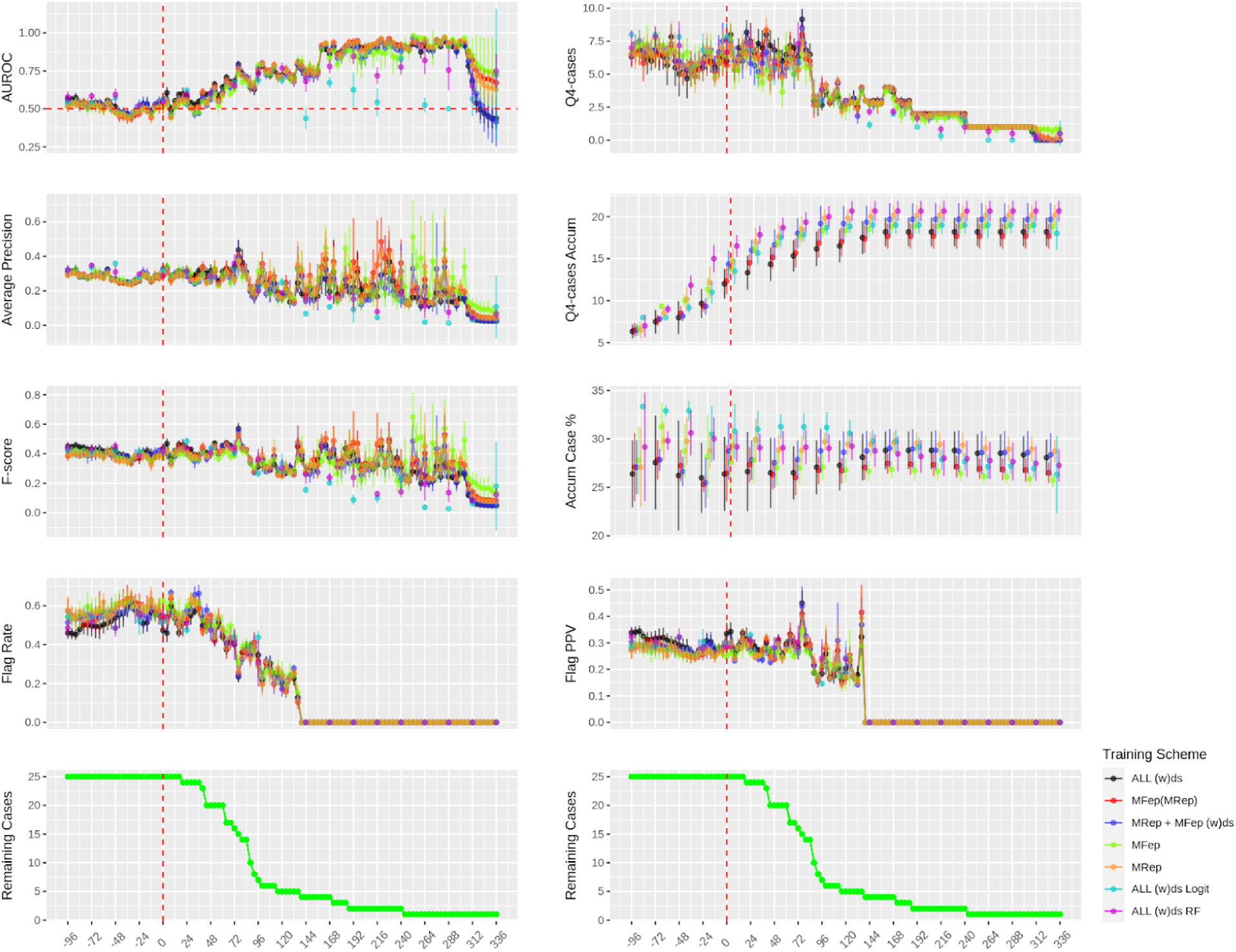
EPIC transferability from *MR_ep_* to *MF_ep_* evaluated by timestep-specific analysis. All predictions made on 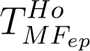 . **ALL**: instance transfer from 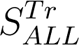 ; **MF_ep_(MR_ep_)**: parameter transfer from 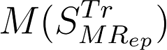 ; **MR_ep_ + MF_ep_**: instance transfer from *MR_ep_*; **MF_ep_**: local learning; **MR_ep_**: brutal-force transfer from 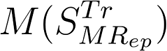 ; **ALL Logit**: Logit based on 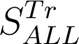 ; **MR_ep_ Logit**: brutal-force transfer with Logit trained on 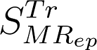 ; **MF_ep_ Logit**: local learning with Logit trained on 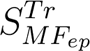

**Fig 5.**
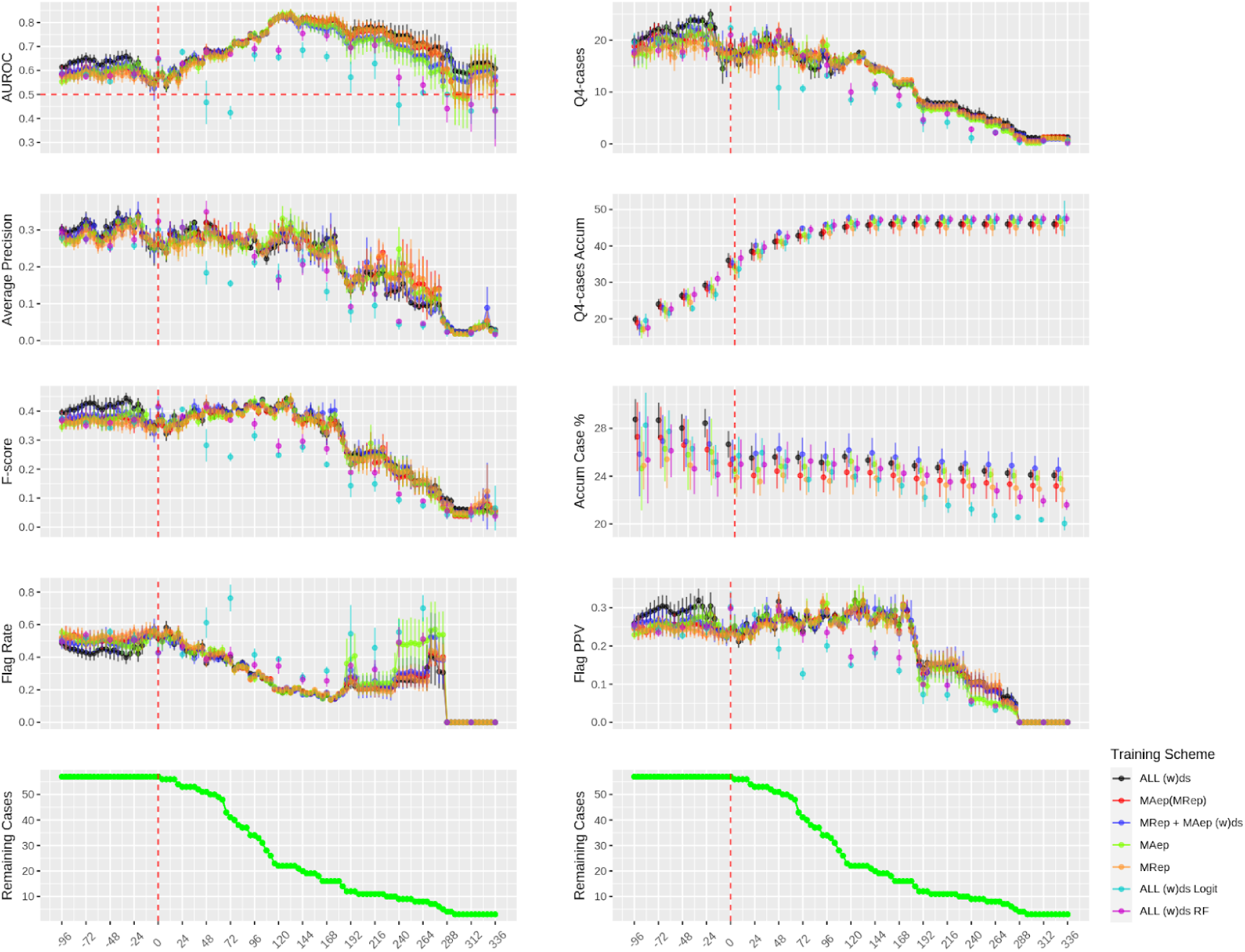
EPIC transferability from *MR_ep_* to *MA_ep_* evaluated by timestep-specific analysis. All predictions made on 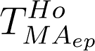 . **ALL**: instance transfer from 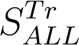 ; **MA_ep_(MR_ep_)**: parameter transfer from 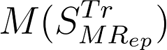 ; **MR_ep_ + MA_ep_**: instance transfer from *MR_ep_*; **MA_ep_**: local learning; **MR_ep_**: brutal-force transfer from 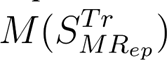 ; **ALL Logit**: Logit based on 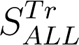 ; **MR_ep_ Logit**: brutal-force transfer with Logit trained on 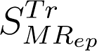 ; **MA_ep_ Logit**: local learning with Logit trained on 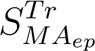

#### Comparison with atemporal models

At various points before the index date and up to three days afterward, the atemporal Logit and RF models markedly surpass the GRU-D models in terms of AUROC, average precision, F-scores, and most notably in Q4-cases and Accum Case % (Fig 3,4 *ALL (w)ds Logit* and *ALL (w)ds RF*). Beyond three days after the index date, the performance of the atemporal models, particularly the Logit models, display significant instability, with performances fluctuating at different timesteps and generally underperform the GRU-D models. Nevertheless, it’s important to note that within the current experimental framework (i.e. conducts daily risk assessments starting four days before surgery), both Logit and RF models identified a significantly larger proportion of ileus patients in the highest risk quartile in two EPIC sites (*MR_ep_*, *MF_ep_*).

Examining the regression coefficients of Logit models over the follow-up period reveals a transition in the primary contributing factors. Factors such as surgery type, hospital site, ileus medication, and smoking status are most influential at 4, 3, and 2 days before the index date. This shifts to skin condition and surgery type as the main factors 1 day before the index date, and further shifts to pain location, dressing type, assisted living status, urine condition, and muscle color on the index date (i.e. immediately after surgery).

### Transferability of model explainability

To explore how model explainability changes under various transfer schemes, we performed permutation feature importance tests on selected scenarios detailed below. a) Brutal-force transfer from Centricity to EPIC within site (Fig 6 a). b) Instance transfer between Centricity and EPIC within site (Fig 6 b). c) Brutal-force transfer between EPIC of different sites (Fig 6 c). d) Instance transfer between EPIC of different sites (Fig 6 d).

**Fig 6.**
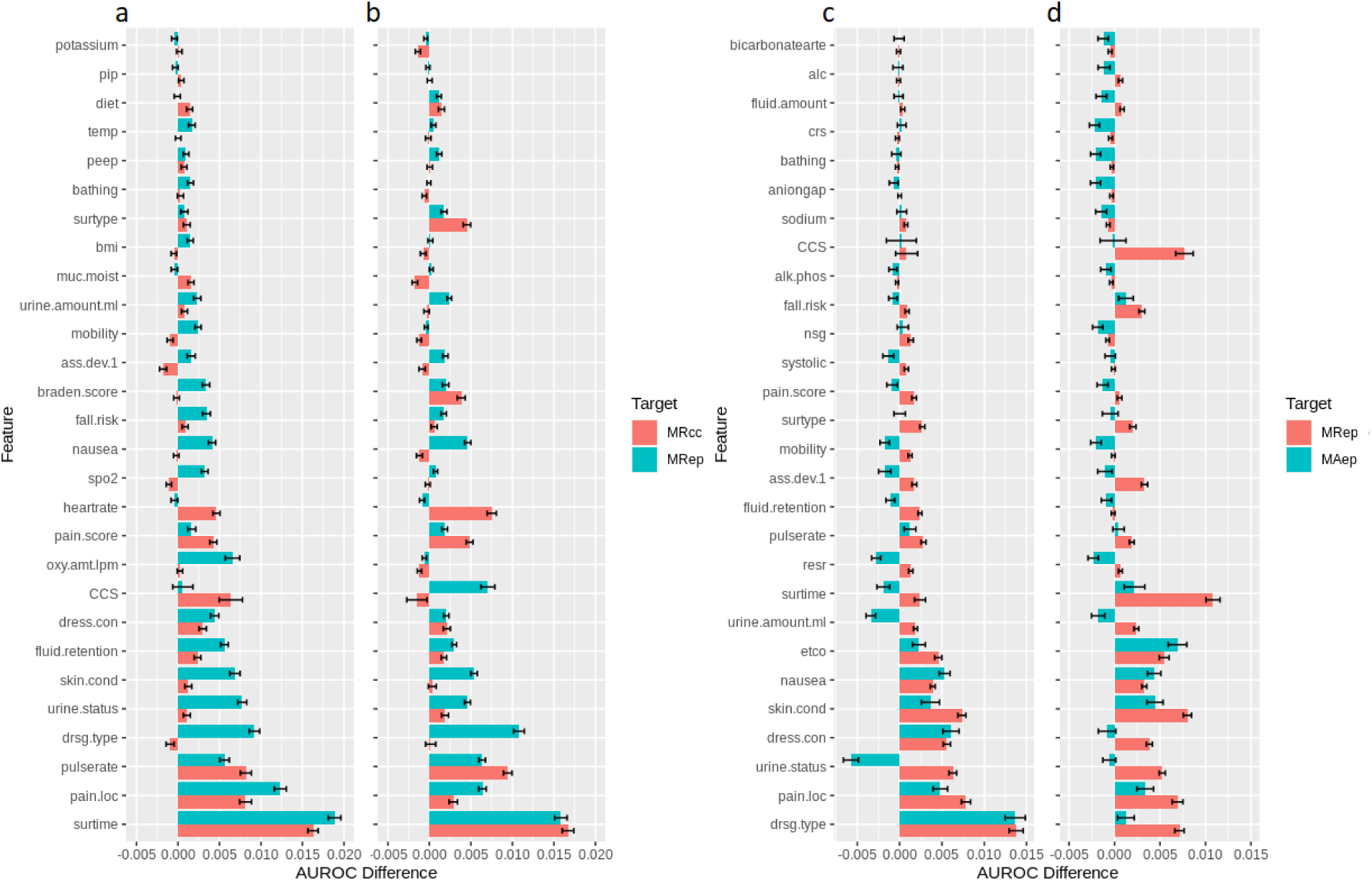
Permutation feature importance test of different transferring scenarios. a) 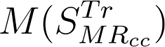 explain 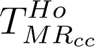 vs 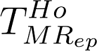. b) 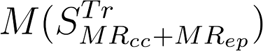 explain 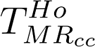 vs 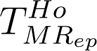. c) 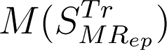 explain 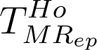 vs 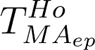. d) 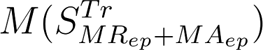. explain 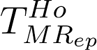 vs 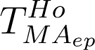. For each pair of permutation tests, the top 20 features were retrieved and merged to generate a final list.

In brutal-force transfer from Centricity to EPIC system within hospital site (Scenario a), we observed 12 overlapping features among top 20 features from each target dataset. The features’ importance varies widely after brutal force transfer (Fig 6 a). Interestingly, instance transfer (Scenario b) resulted in a feature importance pattern remarkably similar to that of brutal-force transfer (Fig 6 b vs a), with the exception for a few features like CCS code and oxygen amount.

In brutal-force transfer between EPIC systems across hospital sites (Scenario c), several features (e.g. urine status, urine amount, surgery time) have no contribution when brutal-force transferred to another hospital site (Fig 6 c). Instance transfer (Scenario d) modified explanation to a subset of features (e.g. CCS, surgery time, dressing condition/type) while maintaining the significance of several key attributes (e.g. pain location, urine status, skin condition, nausea) largely in line with that of brutal force transfer.

### ICD diagnosis date versus ICD post date

In clinical practice, there is often a delay of 1-4 days before the ICD diagnosis date becomes available in the electronic health record (EHR) system. As a result, the ICD post date is more readily accessible for integration into risk models. To evaluate the impact, we trained models using multi-source instance transfer and incorporated dynamic ICD diagnosis date, dynamic ICD post date, and static ICD post date (i.e. use whatever ICD code available 4 days before surgery and replicated through follow-up) as predictors for predicting the hell-out data of MRep, MFep, and MAep, respectively. In our analysis (Supp Fig 2), we observed no significant difference in performance when using these different dates.

## Discussion

Over the past decade, extensive research has revolved around the integration of AI into healthcare, but only a handful of AI tools have undergone thorough validation, and even fewer have been put into clinical practice ^25^ ^26^ ^27^ ^28^. An important part of this challenge lies the lack of generalizability and transferability research with large scale clinical data. Here we provide compelling evidence of the transferability of GRU-D architecture in predicting POI at multiple time points of follow-up across EHR systems and hospital sites. Our findings align with previously reported transferability of RNN based RETAIN models in predicting heart failure across hospitals ^29^, which however only reported static risk prediction. These outcomes support the potential applicability of such models in improving clinical predictive tasks across diverse healthcare environments.

Despite the extreme sparsity in the input feature space, brutal-force transfer maintained remarkably consistent performance across EHR systems and hospital sites, maintaining reasonable stability in the explanation of feature importance. This leads to two intriguing insights: 1) The ground truth behind input features, informative missing, and POI outcome is embedded within each local dataset, which the GRU-D architecture has managed to capture to a certain degree. 2) The contribution of each feature to the outcome is intrinsic to the dataset and less relevant to the model training process. Whereas the variation in how the model explains certain features (such as CCS codes, surgery time, dressing condition/type) suggests that incorporating data from other instances enables the model to find a more effective pathway to predict outcomes.

In essence, instance or parameter transfers offer only a slight advantage over local learning or brutal-force transfer when it comes to the direct improvement of evaluation metrics. However, they notably outperform in terms of variance reduction, particularly in scenarios where the available training data is extremely limited. This reduction in variance leads to predictions that are more precise and therefore more useful for supporting interventions. Furthermore, these findings underscore the ability of GRU-D to effectively handle small datasets with remarkable sparse features. They also imply that for hospitals with restricted access to samples, employing multi-fold cross-validation and averaging the results presents a feasible strategy for applying GRU-D in dynamic risk prediction, despite greater variance.

Negative transfer was observed when transfer from Centricity to EPIC but not the reverse. This could be influenced by 1) the differences in feature distribution, stemming from the non-overlapping colorectal surgery period (Centricity used before 2018, and EPIC after 2018), and 2) variations in how different EHR systems record data (e.g. the records from Centricity system has remarkably more lab and less vital measurements than EPIC system). These findings are consistent with a previous study on transfer learning of CNN-based time series classification ^30^, which indicated that transfer learning is more effective when the source data bears greater similarity to the target data.

Experiments of mutual transfer between the Centricity and EPIC systems reveal that the model performance is primarily influenced by the difficulty of the prediction task, specifically the case prevalence, despite our efforts to address case imbalance through inverse weighting. In contrast, the contribution of the training data on which the model is built and the training or transferring strategy is comparatively less significant. It is anticipatable that for cohorts with extremely rare cases, even the implementation of intricately designed transferring approaches may result in limited improvements in performance. This highlights the challenge of effectively leveraging transfer learning in situations where the target task involves highly uncommon outcomes.

Aligned with our previous findings on superficial infections and bleeding, Logit and RF models demonstrate top-tier performance at specific moments before or immediately after the index date, significantly contributing to the Q4-case related metrics in two hospital sites. Intriguingly, during most of the post-surgical hours, when more current measurements are available, the advantage of atemporal models does not persist and instead exhibits a high level of instability. This counterintuitive performance leads to the following hypotheses: 1) The relatively simple logical structure of static models is insufficient to handle the complexity of features in post-surgical hours. 2) Temporal information plays a crucial role in determining outcomes, which is not captured by static models. 3) The healthcare system’s reliance on a similar static modeling strategy for triggering complication alerts (e.g., bleeding determined algorithmically through hemoglobin levels) may influence results. Despite these observations, we do not dismiss the potential superiority of static models under certain conditions. However, the feasibility of constructing and managing multiple static models deserves further discussion if dynamic risk update is a crucial component.

## Limitations

In this study we focused on the transferability across EHR systems and hospital sites, and didn’t evaluate the data inequality and data distribution discrepancies between racial/ethnic minorities and the medically underserved groups. In our previous studies^31^ ^21^, we found that the EHR system itself significantly impacts the structure and format of clinical data. This influence arises from built-in documentation functionality, such as templates, copy and paste, auto-documentation, and transcription, which can impact the EHR’s specific syntactic and semantic definitions for the data it contains. Additionally, changes in clinical and billing processes, as well as documentation guidelines, may also contribute to this heterogeneity. In the context of PSC-related clinical concepts documentation (e.g., abscess, anemia, purulent drainage, and wound infection), we discovered high syntactic variation and a moderate difference in semantic type and frequency across document sections ^21^. These aforementioned patterns may apply to the ileus documentation, contributing to an increasing number of cases after EPIC migration. A follow-up study is needed to systematically examine this pattern to ensure data quality and process transparency.

## Data Availability

All data produced in the present study are available in de-identified format upon reasonable request to the authors

## Acknowledgement

This work was funded by National Institute of Biomedical Imaging and Bioengineering grant (R01 EB019403) and Cancer Prevention and Research Institute of Texas Established Investigator Award (RR230020).

## Notes

### Competing Interest Statement

The authors have declared no competing interest.

### Author Declarations

The institutional review board of Mayo Clinic gave ethical approval for this work (IRB number: 15-000105).

