## Supplementary Figures for "Revolutionizing Postoperative Ileus Monitoring: Exploring GRU-D’s Real-Time Capabilities and Cross-Hospital Transferability"

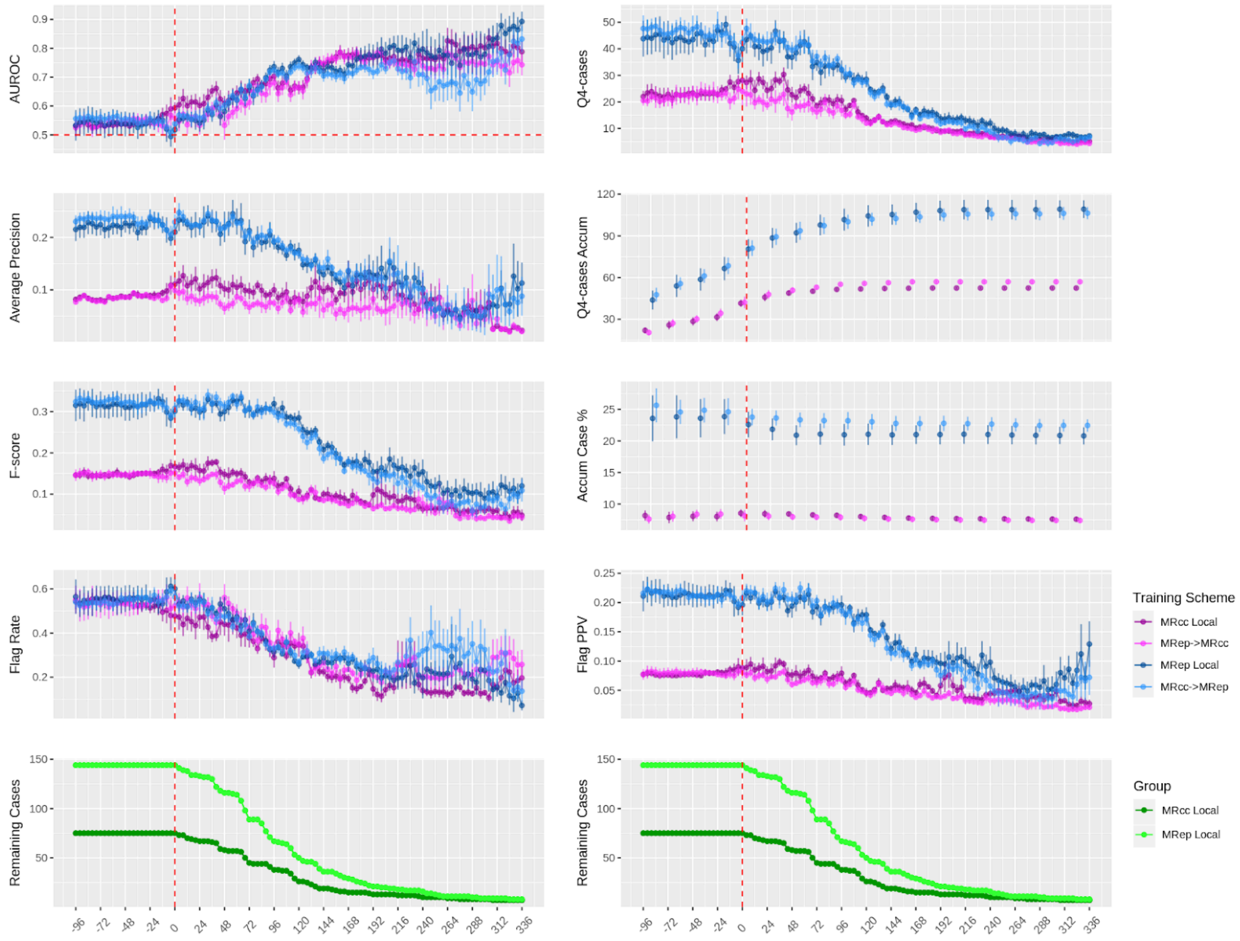


**Sup Fig 1** Brutal-force transfer between $MR_{cc}$ and $MR_{ep}$. **$MR_{cc}$ Local**: Training with $S^{Tr}_{MR_{cc}}$ and apply to $T^{Ho}_{MR_{cc}}$. **$MR_{ep}$->$MR_{cc}$**: Training with $S^{Tr}_{MR_{ep}}$ and apply to $T^{Ho}_{MR_{cc}}$. **MRep Local**: Training with $S^{Tr}_{MR_{ep}}$ and apply to $T^{Ho}_{MR_{ep}}$. **MRcc->MRep**: Training with $S^{Tr}_{MR_{cc}}$ and apply to $T^{Ho}_{MR_{ep}}$

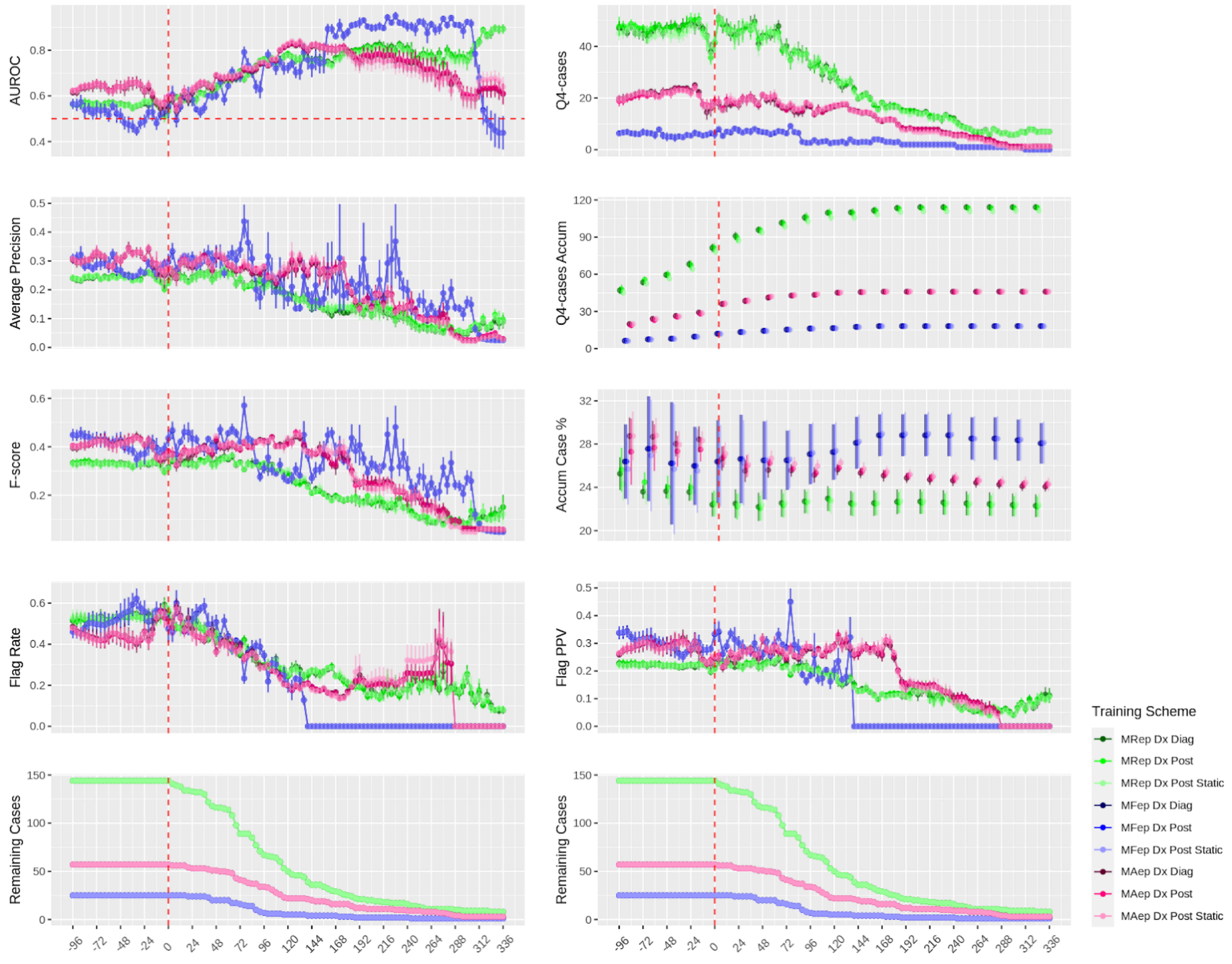


**Sup Fig 2** Comparison of predictions made with dynamic ICD diagnosis date (Dx Diag), dynamic ICD post date (Dx Post), and static ICD post date (Dx Post Static). All models trained with $S_{ALL}^{Tr}$ and dynamic ICD diagnosis date.

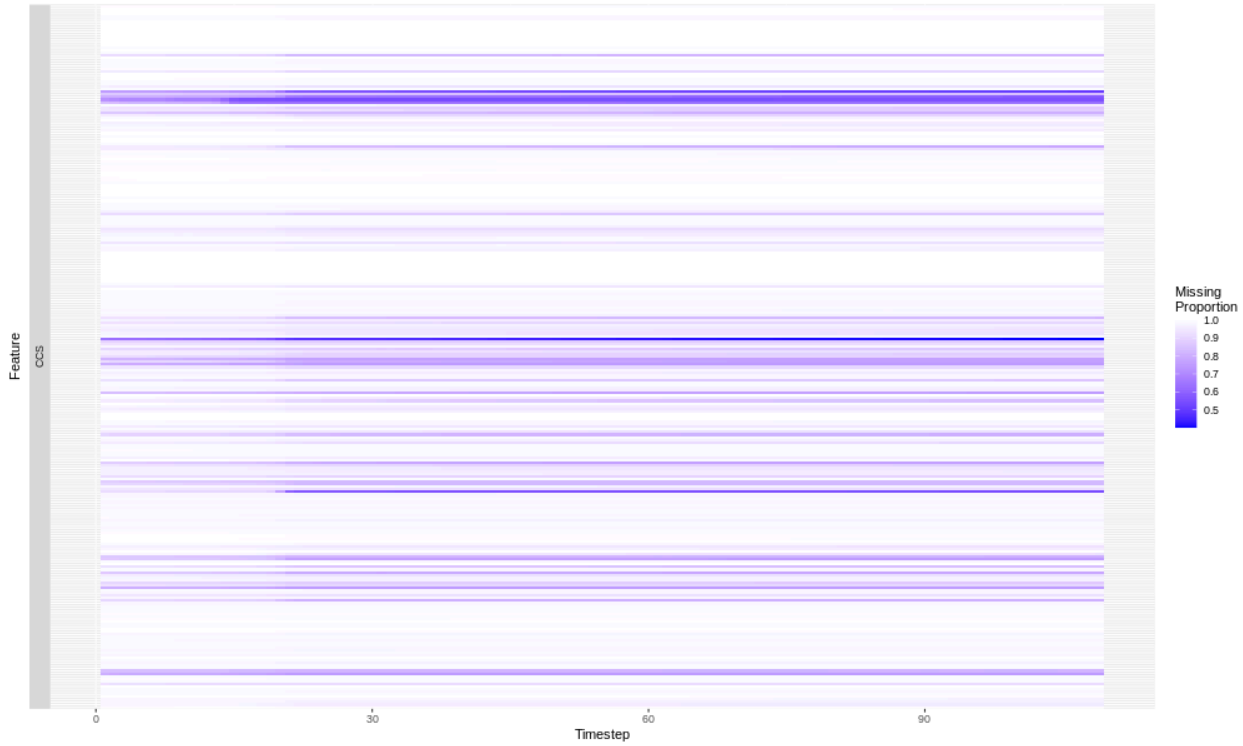


**Sup Fig 3** Missing proportion of 271 CCS codes
